# Health-related quality of life and adherence to physical activity and screen time recommendations in schoolchildren: longitudinal cohort Ciao Corona

**DOI:** 10.1101/2023.03.27.23287611

**Authors:** Sarah R Haile, Samuel Gunz, Gabriela P Peralta, Agnė Ulytė, Alessia Raineri, Sonja Rueegg, Viktoriia Yasenok, Thomas Radtke, Milo A Puhan, Susi Kriemler

## Abstract

**Objectives:** We investigated changes in adherence to physical activity (PA) and screen time (ST) recommendations of children and adolescents throughout the pandemic, and their association with health-related quality of life (HRQOL).

**Methods:** 1769 primary (PS, grades 1-6) and secondary (SS, 7-9) school children from Ciao Corona, a school-based cohort study in Zurich, Switzerland, with 5 questionnaires 2020-2022. HRQOL was assessed using the KINDL questionnaire. PA (≥ 60 min/day moderate-to-vigorous PA) and ST (≤ 2 hours/day ST) recommendations followed WHO guidelines.

**Results:** Adherence to PA recommendations dropped in 2020 (83% to 59% PS, 77% to 52% SS), but returned to pre-pandemic levels by 2022 (79%, 66%). Fewer children met ST recommendations in 2020 (74% PS, 29% SS) and 2021 (82%, 37%) than pre-pandemic (95%, 68%). HRQOL decreased 3 points between 2020 and 2022, and was 9.7 points higher (95% CI 3.0-16.3) in March 2021 in children who met both versus no recommendations.

**Conclusions:** Adherence to WHO guidelines on PA and ST during the pandemic had a consistent association with HRQOL despite longitudinal changes in behavior.

## 1 Introduction

Well-being in children and adolescents is known to be associated with lifestyle factors such as physical activity and screen time [1, 2].

The combination of high physical activity and low screen time has been shown to have a positive and dose-dependent association with well-being in adolescents [3]. The COVID-19 pandemic has been shown to affect children’s quality of life [4, 5, 6, 7, 8]. Further, restrictions related to the early phase of the pandemic were associated with more ST and less PA in children [4, 9, 10], as well with an adverse effect on mental health [11, 12].

Several studies have examined the association between lifestyle and health-related quality of life in children during early phase of the COVID-19 pandemic [6, 7, 13, 14, 15]. Many, however, did not include longitudinal changes, or had a small sample size. Our previous analysis of physical activity, screen time and sleep in children from four cohorts also covered only the first phase of the pandemic [15]. However, public health measures changed much since spring 2021 and after the introduction of vaccines. In Switzerland, public life went almost back to normal in 2021 even though there were still periods of high incidence of SARS-CoV-2 infections, in particular when the Omicron variant started to dominate by the end of 2021.

In Switzerland, public life went almost back to normal since spring 2021, even though there were still periods of high incidence of SARS-CoV-2 infections, in particular when the Omicron variant started to dominate by the end of 2021. Here, our aim was to describe changes in adherence to physical activity and screen time in the Ciao Corona study, a large and longitudinal cohort of children aged 6-16 years from the canton of Zurich in Switzerland over a two year period from June 2020 to July 2022, to examine changes in HRQOL over time. andto examine the hypothesis that adherence to physical activity and screen time recommendations was associated with better HRQOL over time. We hypothesized that adherence to physical activity and screen time recommendations was associated with better HRQOL.

## 2 Methods

### 2.1 Study design, procedures, and participants

The data for this analysis come from the school-based longitudinal cohort study Ciao Corona [16], in which 55 randomly selected schools (primary school grades 1-6 and secondary school grades 7-9, ages 6-17 years) in the canton of Zurich, the largest canton in Switzerland of approximately 1.5 million inhabitants (18% of the total Swiss population), took part. In each of the randomly selected schools, a sample of classes was taken, and all students from those classes were invited to participate. Serological testing was performed at 5 different timepoints from June 2020 to July 2022. Children (or their parents) were also asked to fill out a baseline questionnaire at the time of their first antibody test which was shortly after the lockdown, and then they completed follow-up questionnaires on a periodic basis (July 2020, January 2021, March 2021, September 2021, and July 2022). Schools and classes participating in June 2020 were also invited to participate in later rounds of testing. Therefore, many children in the cohort participated at multiple timepoints.

Ciao Corona was approved by the ethical committee of the canton of Zurich (2020-01336), and the study design has been published elsewhere [16] (ClinicalTrials.gov identifier: NCT04448717). All participants provided written informed consent before being enrolled in the study. Serological results from June 2020 through December 2021 [17, 18] as well as changes in lifestyle behaviors through April 2021 [15] have been reported previously.

We had data on 1875 to 2500 children and adolescents (hereafter, children) assessed at each of 5 timepoints. In order to ensure a longitudinal cohort with repeated online questionnaires for each child, we restricted our sample to children and adolescents who had completed a baseline questionnaire in June 2020 and at least one follow-up questionnaire.

For context, pandemic restrictions in Switzerland included a brief lockdown from March to May 2020 during which schools were completely remote [19], with a partial return to inperson school (e.g. with smaller classes or hybrid in person and online) until July 2020 and full in-person attendance thereafter. Despite having quite high case rates [20], schoolchildren in Switzerland remained at school with additional regional regulations which included mask wearing (primary school from Jan. 21, 2021 for 4th graders and above or Dec. 9, 2021 for 1st graders and older to Feb. 20, 2022, secondary school from Oct. 28, 2020 to Feb. 20, 2022), quarantine and isolation measures (10 day requirement until Jan. 12, 2022, then 5 day requirement until March 30, 2022) and weekly pooled testing in some, but not all schools (from August 2021 to Feb. 20, 2022). Other activities, such as extracurricular sports, continued with the use of masks, small groups, availability only to children under the age of 16, and other restrictions through March 2021. Thereafter, most activities both in and outside of school continued without restrictions for children and adolescents. The situation in Switzerland during the pandemic provides an opportunity to assess changes in lifestyle and HRQOL where preventive measures for children were relatively mild compared to other countries [21].

### 2.2 Outcomes and Exposures

The outcomes considered in this analysis were health-related quality of life (HRQOL), as well as self-rated health and life satisfaction. HRQOL was measured using the KINDL questionnaire [22], a validated measure of HRQOL in children comprising 24 questions on a Likert scale, which are converted to a total score from 0 (worst) to 100 (best). The KINDL questionnaire has 6 subscales: physical, emotional, self-esteem, family, friends, and school (each also on a scale from 0 to 100). For self-rated health [23], participants were asked to categorize their overall health status as excellent, good, reasonably good, or bad. For life satisfaction, participants were asked to rate their life on an 11-point scale of 0 (worst) - 10 (best) using the Cantril ladder [24]. Along with the above mentioned timepoints, children were asked about pre-pandemic levels of PA and ST, but not about previous HRQOL.

The exposures of interest were physical activity and screen time. Physical activity was assessed by asking how many hours during the week, and on weekends, participants spent on physical activity (with light sweating). The average time spent on physical activity was then computed across the entire week. If only one of those questions was answered, we assumed that this information applied to the entire week. For screen time, participants were asked how many hours per week they spend with electronic media (e.g. smartphone, tablet, computer, television, or gaming), not including media use in school. Exact wording of questions is found in the supplementary material. Continuous measures of physical activity and screen time were converted to categorical variables denoting whether they met international recommendations [25, 26]: at least 1 hr/day of physical activity, and no more than 2hr/day of screen time. A combined exposure to both behaviors was considered as a variable with 4 levels: did not meet either recommendation, met only physical activity recommendations, met only screen time recommendations, and met both physical activity and screen time recommendations.

Covariates, collected from the previously mentioned questionnaires, included participants’ age and sex as collected in June 2020, as well as body mass index (BMI) Z-score at baseline [27] and presence of chronic conditions. Age was divided into two groups, those who remained in primary school during the entire timeframe (1st through 6th grades, approximately ages 6-10 years at the beginning of the pandemic period), or those who were in secondary or entered secondary school during the pandemic (ages 11-16). In the baseline questionnaires, information on parents’ nationality (at least one parent Swiss, vs both non-Swiss) and education level (at least one parent with preparatory high school or university vs apprenticeship or professional school) was collected. Children were considered overweight if their baseline body mass index (BMI) age and sex adjusted Z-score was 1 or higher. Participants were asked about diagnoses of the following chronic conditions: asthma, hay fever, diabetes, inflammatory bowel disease, ADHD, epilepsy, depression / anxiety and joint disorders. Other conditions that were included on the questionnaire but not counted as chronic conditions for this study were: celiac disease, lactose intolerance, other allergies, neurodermatitis/excema, and other diseases with a comment field. None of the participants in this analysis reported other conditions that were not already listed.

### 2.3 Statistical Methods

Changes in physical activity, screen time and HRQOL were examined graphically, and using tables by summarizing as n (%) or median and interquartile range (IQR). To assess the association of physical activity or screen time or both with HRQOL at each time point, we used inverse probability weighting with propensity scores which adjusted for the probability that the participant had the observed physical activity or screen time adherence status given all previous covariates (described above) and all previous lifestyle and HRQOL measurements [28]. Missing data were imputed using multiple imputation [29] prior to fitting the inverse probability weighted models (25 imputations). As a sensitivity analysis, we also considered models that were stratified by age group (primary vs secondary school). Interpretation of the model results depended on the clinical significance of any observed effects and their confidence intervals, and not on p-values [30].

The statistical analysis was conducted using R (R version 4.2.1) [31]. Multiple imputation was performed using the mice [32], while inverse probability weights were computed using the Weightit [33]. The models were then fit using generalized estimating equations with the geepack package [34]. Plots were created using the ggplot2 package [35].

## 3 Results

Among the 1769 children and adolescents included in this analysis (Table 1), 1126 (64%) remained in primary school for all of the study questionnaires, 52% were male, and 11% were overweight. 85% of participants had at least one Swiss parent, and 75% reported having at least one parent with a high level of education. 20% of participants reported having a chronic condition which potentially interferes with lifestyle or HRQOL (see Table S1).

**Table 1:**
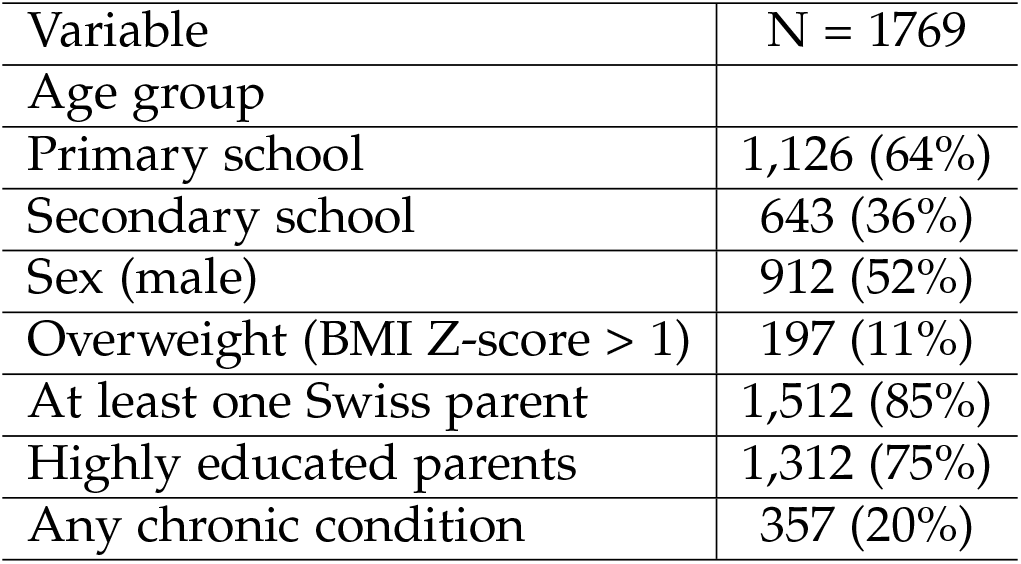
Baseline characteristics. Lifestyle and health-related quality of life analysis of Ciao Corona, Switzerland, 2020 - 2022. Primary school includes grades 1-6, while secondary school is grades 7-9. Chronic conditions included hay fever (14%), attention deficit hyperactivity disorder (ADHD, 4%), asthma (4%), depression (0.3%), epilepsy (0.2%), joint disorders (0.2%), diabetes (0.1%) and inflammatory bowel disease (<0.1%) (Table S1).

### 3.1 Adherence to physical activity and screen time recommendations

Figure 1 shows changes in adherence to physical activity and screen time recommendations over time, by age group (see also Table S2). Participants in secondary school generally had less physical activity and more screen time than those in primary school (for example 77% secondary vs 83% primary at baseline), especially during the lockdown period (52% secondary vs 59% primary), and greater differences in screen time than in physical activity were observed between the age groups (for example 68% secondary vs 95% primary at baseline). In the early phases of the pandemic, participants were less physically active (in July 2020, 59% primary, 52% secondary adhered to physical activity recommendations) and had more screen time (74% primary, 29% secondary adhered to screen time recommendations) than pre-pandemic (83% primary and 77% secondary for physical activity, and 95% primary and 68% secondary for screen time). By early 2021, screen time had decreased back to almost pre-pandemic levels (93% primary and 53% secondary), but has slightly increased again since then (in July 2022, 82% primary and 38% secondary). The percentage of children meeting physical activity recommendations remained low through early 2021 (in March 2021, 75% primary and 87% secondary), but then increased and remained at pre-pandemic levels.

**Figure 1:**
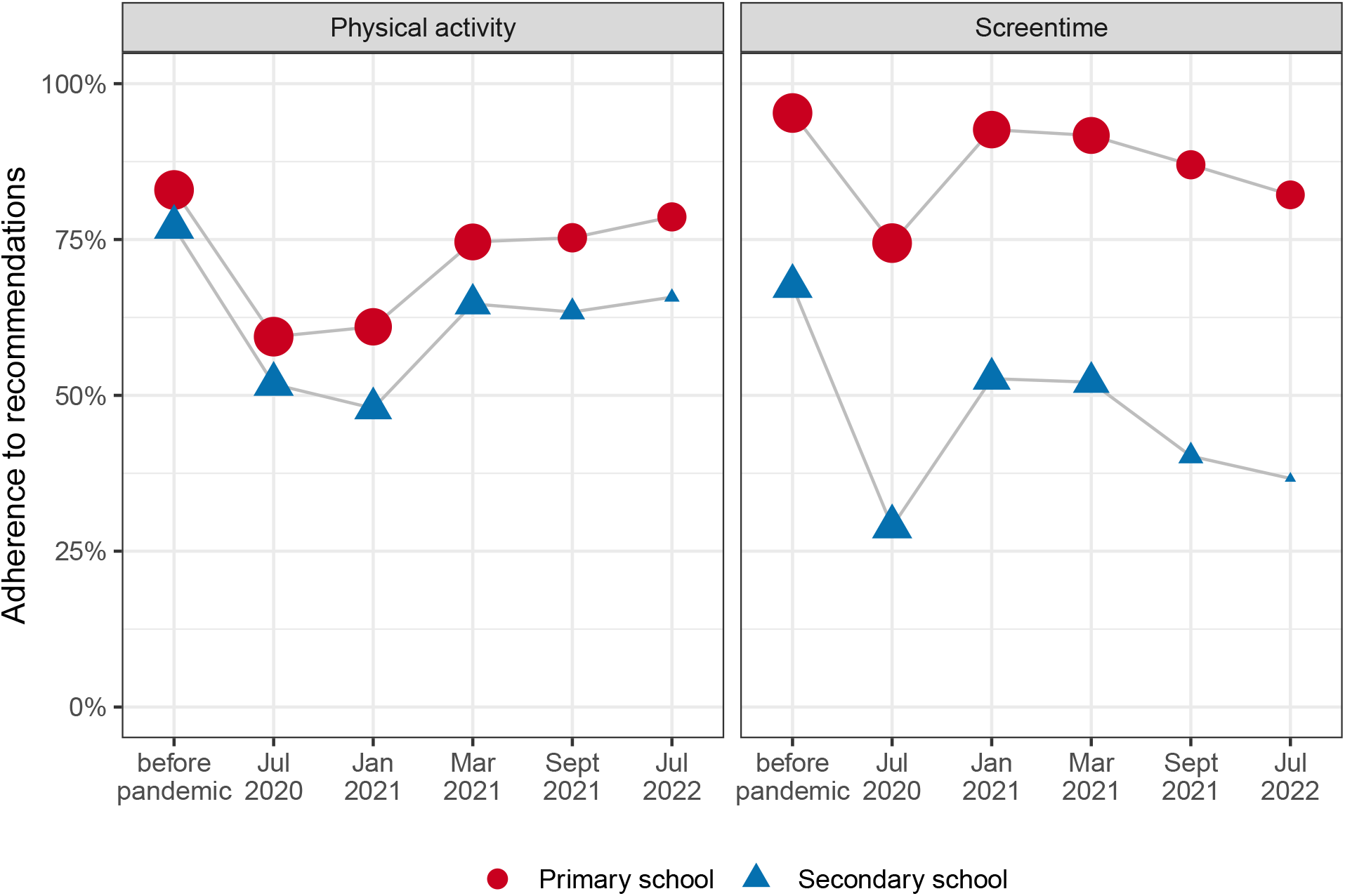
Adherence to physical activity and screen time recommendations during the COVID-19 pandemic, by age group. Point size corresponds to the number of observations. Ciao Corona, Switzerland, 2020 - 2022.

### 3.2 Longitudinal changes in HRQOL

HRQOL, as measured by KINDL total score, was similar across all timepoints (median 77, IQR 71 - 84), but highly variable across the sample (range 17 to 100), and reduced approximately 3 points between July 2020 and July 2022 (Figure 2, Table S3). Children not meeting either physical activity or screen time recommendations had on average lower HRQOL outcomes than those meeting physical activity or screen time or both recommendations. This pattern was consistent across all follow-up questionnaires.

**Figure 2:**
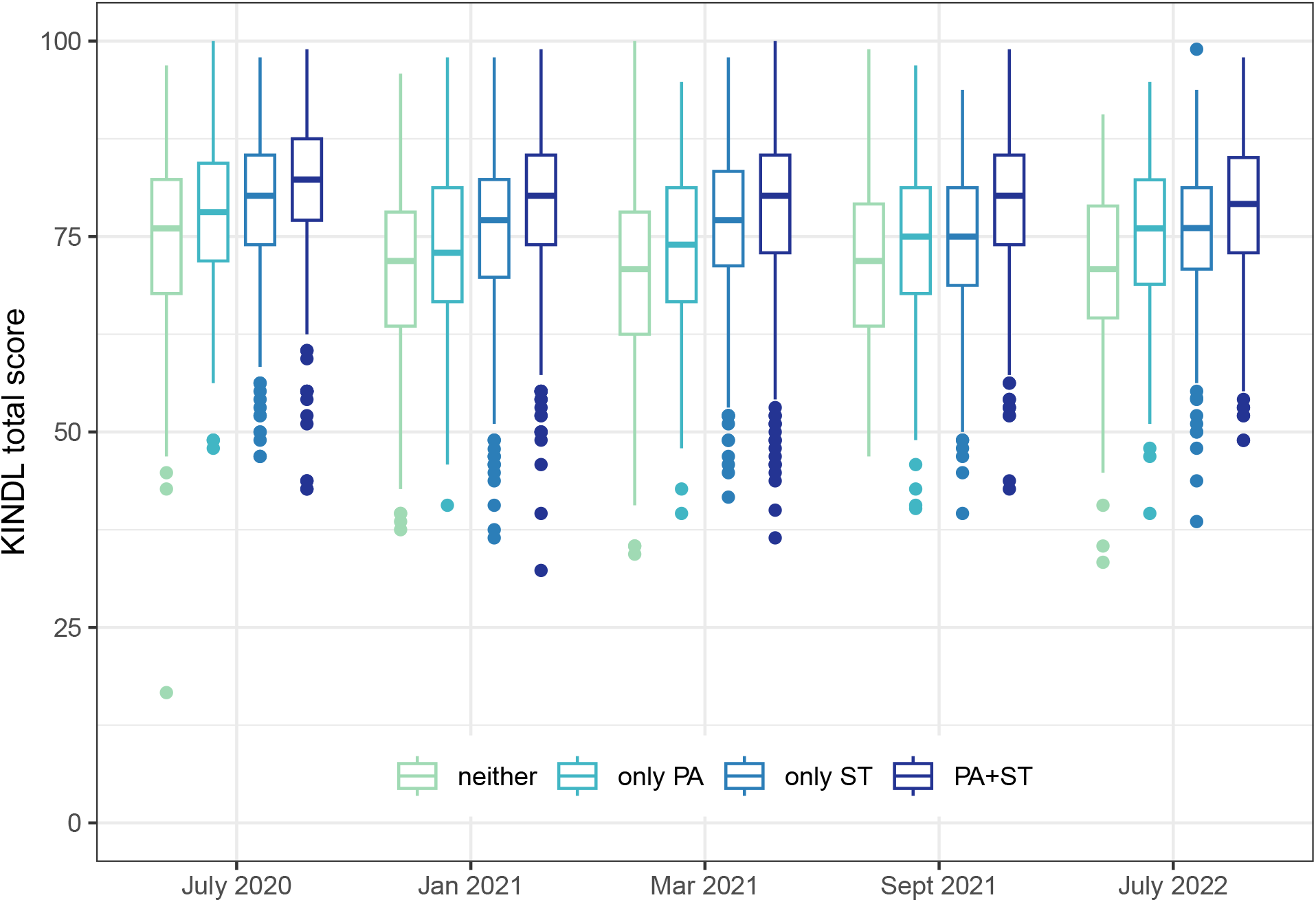
Health-related quality of life over time, by adherence to physical activity and screen time recommendations. Adherence to recommendations had the reference category of not adhering to either recommendation (“neither”), compared to only adhering to physical activity recommendations (“only PA”), only adhering to screen time recommendations (“only ST”) and adhering to both recommendations (“PA+ST”). Ciao Corona, Switzerland, 2020 - 2022.

### 3.3 Association between HRQOL and physical activity and/or screen time

Participants meeting both physical activity and screen time recommendations had 2.4 (July 2022, 95% CI -1.3 to 6.1) to 9.4 (March 2021, 3.0 to 16.3) points higher total KINDL scores than those not meeting either of the recommendations (Figure 3, Table S4). Children meeting screen time but not physical activity recommendations had 0.5 (July 2022, 95% CI -3.4 to 4.4) to 8.1 (March 2021, 1.1 - 15.2) points higher KINDL scores than those who did not meet either recommendation, while those meeting physical activity but not screen time recommendations had 1.7 (July 2022, -2.3 to 5.6) to 6.2 (March 2021, -0.5 to 12.9) point higher KINDL scores. The magnitude of the observed differences due to meeting both physical activity and screen time recommendations compared to meeting neither of the recommendations increased from June 2020 (4.6 points, 95% CI 2.8 - 6.5) to March 2021 (9.7, 3.0 to 16.3), after which the association decreased to 2.4 (−1.3 to 6.1) points. A similar pattern was seen in each of the KINDL subscales (Figure S2, Tables S5 and S6), life satisfaction and self-rated health (Figure S3 and Table S7). We also observed similar patterns in an age-stratified analysis, but do not present those results due to the relatively small sample size (Table S8).

**Figure 3:**
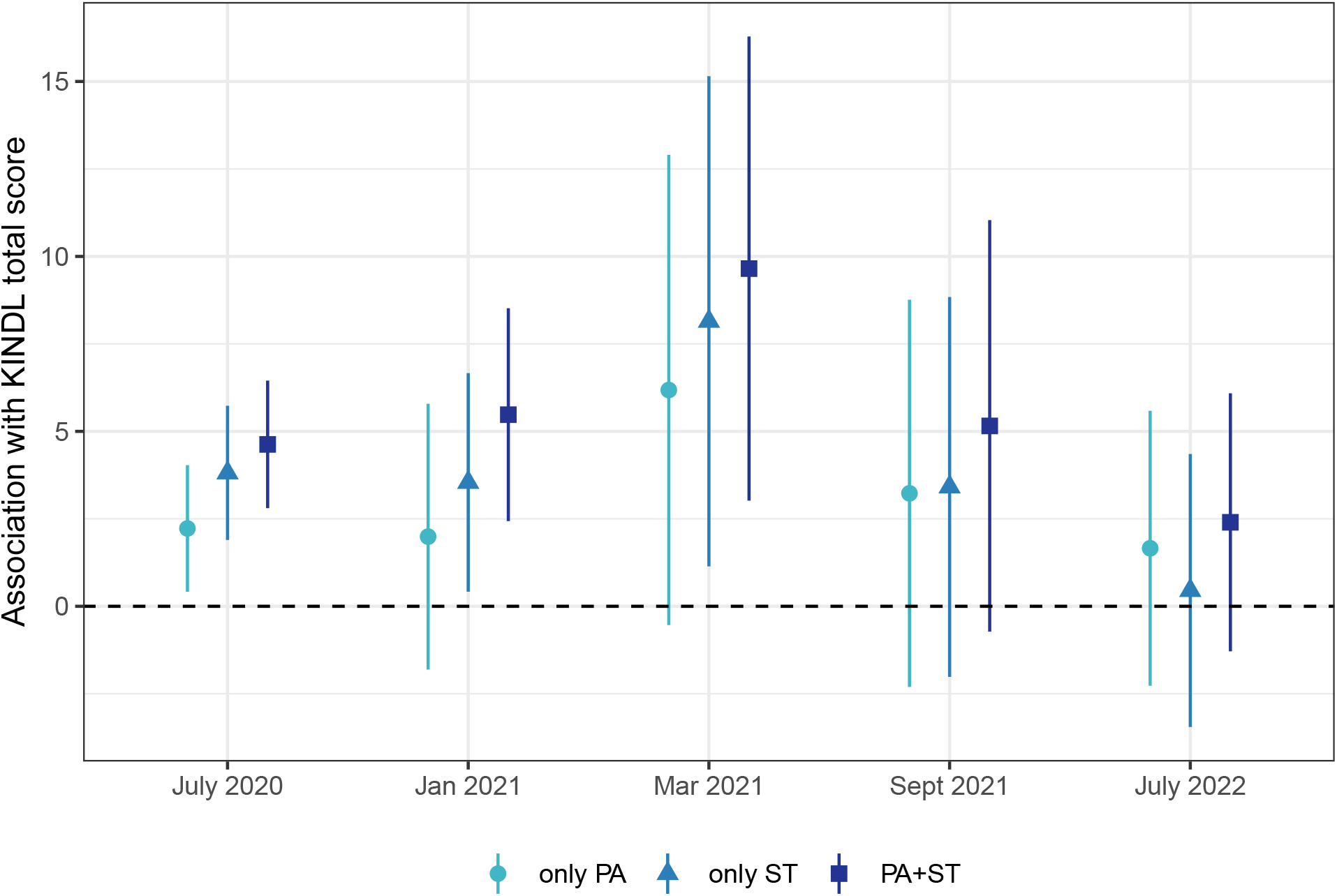
Adjusted association between health-related quality of life and adherence to physical activity (PA) and screen time (ST) recommendations, fit using inverse probability weighted models and adjusting for past covariates (age, sex, nationality, parents’ education, presence of chronic conditions, and body mass index) and previous adherence to recommendations. Adherence to recommendations had the reference category of not adhering to either recommendation, compared to only adhering to physical activity recommendations (“only PA”), only adhering to screen time recommendations (“only ST”) and adhering to both recommendations (“PA+ST”). Ciao Corona, Switzerland, 2020 - 2022.

## 4 Discussion

In this longitudinal cohort study, we observed that fewer children met physical activity and screen time recommendations during a short lockdown and through early 2021 than in the time periods before or after. Primary school children were much more likely to meet physical activity and screen time recommendations, especially during the prepandemic and lockdown. HRQOL, as measured by total KINDL score, appeared relatively constant among children in both age groups during the course of the COVID-19 pandemic, even though variability among the tested children and adolescents was high, as seen in Figure 2, Table S4). Children meeting both physical activity and screen time recommendations had the highest HRQOL scores on average, followed by those only meeting screen time recommendations, those only meeting physical activity recommendations and then those not meeting either recommendation. After adjusting for past lifestyle behaviors, there was a statistically significant and clinically meaningful difference (ranging from 4.6 points in July 2020 to 9.7 points in March 2021, Table S3) in HRQOL between children meeting and not meeting physical activity and screen time recommendations. This association was consistent despite overall high variability in HRQOL. Nevertheless, the temporal evolution of the association between lifestyle and HRQOL showed a peak in spring 2021 and then reduced in summer 2022.

The main aim of our study was to test the hypothesis that adherence to physical activity or screen time was associated with HRQOL in children and adolescents. Adherence or non-adherence to physical activity and screen time recommendations was associated in our study with a difference of between 0.5 and 9.7 points on the 100-point KINDL HRQOL scale, with 50% of the effects between 2.3 and 5.3 (Figure 3, Table S2). While many features of KINDL have been discussed previously, no minimal important difference has been reported [36, 37]. We searched the literature for comparisons of HRQOL in healthy children compared to children with chronic conditions to explore the range of differences observed. A comparison of children with and without asthma indicated a difference of about 4.8 points [38], while children with headaches had a 3.5 point lower KINDL total score than those without headaches [39]. Other studies have shown differences of 0.8 points in overweight children, 2-4 points in children with hemophilia, and 9 points in children with various chronic conditions when compared to children without such conditions [40, 41, 42]. These results indicate that a difference of 2-5 points as observed in this study are likely to be relevant in practice.

A second objective was to tease out temporal changes of the association between fulfillment of physical activity and screen time recommendations and HRQOL. We observed a consistent and increasing association between HRQOL and lifestyle through March 2021 which then diminished to a weak association by mid 2022, This trend over time was similar to that seen pre-pandemic in other cohorts [43] or to early pandemic data documenting increased life satisfaction with higher physical activity and lower screen time [13, 44, 45]. Our strongest association by March 2021 coincided with the cessation of the multiple pandemic restrictions for the students (Figure S1). It is possible that physical activity and screen time explained more of the variance in HRQOL at that time when many other factors were out of children’s control due to pandemic measures that were still perceived as existent.

The results from our study are in line with the literature. A number of studies have shown reduced physical activity during the COVID-19 pandemic [46, 47], as well as prolonged screen time [9, 11]. We observed a consistent and increasing association between HRQOL and lifestyle through March 2021, which then diminished to a weak association by mid-2022, similar to that seen pre-pandemic [1, 48, 49, 50, 13]. A 2021 review noted that increased satisfaction was associated with higher physical activity and lower screen time [3], while a 2020 cross-sectional survey observed that children with higher physical activity and lower screen time had fewer difficulties and mental health symptoms [48]. The strong association observed in March 2021 may be due to various factors, including possible seasonal variation [51, 52]. More likely however is that the end of multiple pandemic restrictions [19] (Figure S4) led to changes in participants’ behaviors and HRQOL (or perhaps perception of their HRQOL). It may be that more physical activity and screen time explained more of the variance in HRQOL at that time in the context of the pandemic, or that lifestyle played a larger role in determining HRQOL when many other factors were out of children’s control due to pandemic measures. Due to the specific mix and severity of pandemic measures in Switzerland, our results may not necessarily be generalizable to other countries or populations. Additionally, the association between HRQOL and lifestyle may have been stronger in countries with stronger pandemic restrictions which included longer school closures, or in populations with fewer socio-economic resources.

Some implications for public health and further research can be inferred from these results. First, measures to increase physical activity and reduce screen time in children and adolescents are important for both physical health and HRQOL. This may be especially true in times of pandemics where HRQOL as well as lifestyle may be and become restricted by various preventive measures, for example those that disable self-determined and school-based PA, but also many other activities that impact HRQOL such as social interaction with peers. Public health measures to improve health and well-being in children and adolescents should therefore aim to affect both lifestyle and HRQOL from the very beginning of any upcoming pandemic. The more extensive the restrictions take place, the more action by Public health and school authorities should be taken up to prevent the health and HRQOL compromising burden of pandemic-related restrictions. This is especially true for future pandemics that should recognize how big the COVID-19 induced burden on lifestyle, mental health and HRQOL worldwide was for children and adolescents, with our study likely representing the least form of the negative, yet clinically meaningful impact in a country with the mildest form of restrictions and a socio-economically stable population. Although we could not find a more pronounced impact of the pandemic on some subgroups at increased risk for compromised lifestyles and HRQOL even during “out of pandemic” times, a special emphasis should be put on children and adolescents with overweight, chronic health conditions or psycho-social problems [53, 54]. While we examined the relationship to adherence to WHO guidelines for physical activity and screen time, it may well be that a dose-response relationship exists where more physical activity and less screen time leads to higher HRQOL even if recommendations are not met. Dose-dependent relationships in adolescents were found previously [3], but could be the topic of further research in children 10 years old and younger. Second, high variability in HRQOL across our sample indicates that other factors besides lifestyle are likely to play a role in individual HRQOL. Studies exploring possible determinants of HRQOL in children and adolescents, especially during a pandemic like COVID-19, could therefore be of use in adapting measures to increase physical activity and reduce screen time, as well as in identifying individuals that may need additional support in these areas.

The main strength of this study is the use of a large longitudinal sample of children and adolescents assessed repeatedly across 2 years of the COVID-19 pandemic. Physical activity and screen time as key lifestyle factors were included as exposure variables, and we were able to adjust for a large set of relevant confounders (age, sex, BMI, chronic conditions, nationality, parents’ education and school unit). Therefore, if for example, body mass index or parental education were the main drivers of HRQOL during the pandemic, the effects of physical activity and screen time would no longer have been large enough to be considered relevant. The use of inverse probability weighting allowed to estimate a causal effect of physical activity and screen time on HRQOL while adjusting for past confounders and past lifestyle. Limitations of this study include lack of HRQOL data prior to the pandemic, and recall bias as HRQOL, physical activity and screen time were all assessed subjectively using questionnaires. Despite the lack of objective measurement of physical activity, we observed typical differences in physical levels between children and adolescents. Recall bias should however not affect the strength of the association. It is possible that physical activity, screen time and HRQOL had seasonal changes which we cannot account for [52]. Differential loss to follow-up was likely as children who changed schools or classes, or who went to college preparatory secondary school (“Gymnasium”) during the study period could not continue to participate. As with many studies, this sample had more highly educated parents and was more likely to have Swiss nationality than the target population of all schoolchildren in the canton of Zurich. We have assumed that physical activity and screen time influence HRQOL, but it could instead be that HRQOL influenced physical activity and screen time, or that all have common causes.

In conclusion, changes in physical activity and screen time during the COVID-19 pandemic only temporarily translated to substantial changes in HRQOL, and were minimal by June 2022. Yet, meeting physical activity and / or screen time recommendations was consistently associated with statistically significant and clinically meaningful changes in HRQOL, and the magnitude of those differences likely exceed the minimal important difference. The high variability in HRQOL implies that physical activity and screen time are not the sole driving factors of HRQOL in children, and other factors must also play a role.

## Data Availability

Upon study completion in 2023, de-identified and potentially aggregated participant data, together with required data dictionaries, will be available on reasonable request by email to the corresponding author. The purpose and methods of data analysis will be evaluated by the study team first to ensure that it complies with the ethics approval.

## Author Contributions

SK and MAP initiated the project and preliminary design. SK, TR, SRH developed the study question and methodology. SK, TR, AU, AR and SR recruited study participants, collected, and managed the data. SG supported the organisation and data collection at schools for the last testing round (July 2022). GPP, SG and SRH organized and cleaned the data for the analysis of lifestyle and HRQOL. SRH performed statistical analysis and wrote the first draft of the manuscript. VY performed a review of the literature. All authors contributed to the development of the study question and interpretation of its results and revised and approved the manuscript for intellectual content. SK, SR, AR and SRH had access to and verified all underlying data. The corresponding author SK attests that all listed authors meet authorship criteria and that no others meeting the criteria have been omitted.

## Ethical Approval

The study was reviewed and approved by the Ethics Committee of the canton of Zurich (2020-01336), Switzerland. Written informed consent to participate in this study was provided by the participants’ legal guardian.

## Acknowledgments

We thank Miquel Serra-Buriel for consulting on the statistical methods, and Jan Schlegel for summarizing the pandemic measures.

