## Supplementary material for "Health-related quality of life and adherence to physical activity and screen time recommendations in schoolchildren: longitudinal cohort Ciao Corona": Table S

### List of Figures

S1 COVID-19 restrictions for schoolchildren in the Canton of Zurich January 2020 - July 2022. Home office for the parents was obligatory (complete) or only recommended by the Federal Council (partial). Schools had to be closed (complete) or the timetables were reduced (partial). Sport and other public facilities were closed (complete) or there were only restrictions of sporting activities and gatherings in public areas (partial). Class camps were either forbidden (complete) or could only be conducted in compliance with a protective concept (partial). Finally, during some periods masks were mandatory in school (complete). . . . 3

S2 Model results for association of HRQOL subscales with adherence to physical activity and screen time recommendations (compared to reference category of not adhering to either recommendation). . . . 4

S3 Model results for association of self-rated health and life satisfaction with adherence to physical activity and screen time recommendations (odds ratio (95% confidence interval)) . . . 5

S4 HRQOL by age at baseline . . . 8

Excerpt of questions about physical activity and screen time

1. On a typical weekday (Monday through Friday), how many hours per day does your son/daughter CURRENTLY engage in exercise or other physical activity (that caused sweating or breathing more)? Please count school sports.
2. On a typical weekend day (Saturday, Sunday), how many hours per day does your son/daughter CURRENTLY do sports or other physical activity (that caused sweating or breathing more)?
3. How many hours a day does your son/daughter CURRENTLY spend using electronic devices (e.g. TV, mobile phone, computer, Playstation, Xbox) in their free time on a typical weekday (Monday to Friday)?
4. How many hours a day does your son/daughter CURRENTLY spend using electronic devices (e.g. TV, mobile phone, computer, Playstation, Xbox) in their free time on a typical weekend day (Saturday and Sunday)?

Table S1: Chronic conditions reported. Lifestyle and health-realted quality of life analysis of Ciao Corona, Switzerland, 2020 - 2022.

| Variable | N = 1769 |
|---|---|
| Any chronic condition | 357 (20%) |
| Hay fever | 253 (14%) |
| Attention deficit hyperactivity disorder | 75 (4.2%) |
| Asthma | 71 (4.0%) |
| Depression/Anxiety | 5 (0.3%) |
| Epilepsy | 4 (0.2%) |
| Joint disorder, incl. Arthritis | 4 (0.2%) |
| Diabetes mellitus | 2 (0.1%) |
| Inflammatory bowel disease | 1 (<0.1%) |

Table S2: Percentage of children meeting physical activity (PA) or screen time (ST) recommendations in primary and secondary school, by timepoint (see also Figure 1).

| timept | primary_PA | secondary_PA | primary_ST | secondary_ST |
|---|---|---|---|---|
| baseline | 82.9% (934/1126) | 77.0% (495/643) | 95.3% (1073/1126) | 67.5% (434/643) |
| July 2020 | 59.4% (669/1126) | 51.8% (333/643) | 74.4% (838/1126) | 28.9% (186/643) |
| Jan 2021 | 61.0% (610/1000) | 47.9% (276/576) | 92.6% (930/1004) | 52.7% (305/579) |
| Mar 2021 | 74.6% (723/969) | 64.7% (346/535) | 91.7% (894/975) | 52.1% (284/545) |
| Sept 2021 | 75.3% (469/623) | 63.4% (199/314) | 87.0% (535/615) | 40.3% (124/308) |
| July 2022 | 78.6% (482/613) | 65.8% (144/219) | 82.2% (493/600) | 36.7% (77/210) |

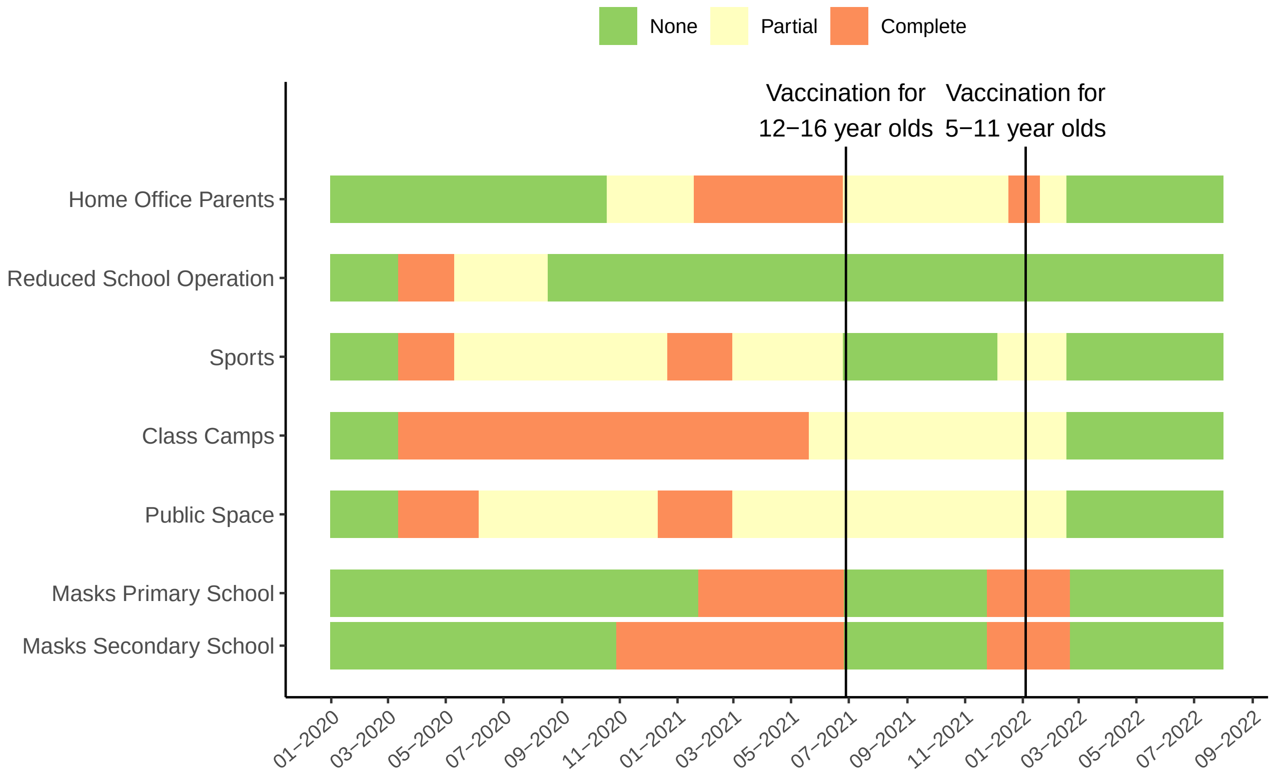

Figure S1: COVID-19 restrictions for schoolchildren in the Canton of Zurich January 2020 - July 2022. Home office for the parents was obligatory (complete) or only recommended by the Federal Council (partial). Schools had to be closed (complete) or the timetables were reduced (partial). Sport and other public facilities were closed (complete) or there were only restrictions of sporting activities and gatherings in public areas (partial). Class camps were either forbidden (complete) or could only be conducted in compliance with a protective concept (partial). Finally, during some periods masks were mandatory in school (complete).

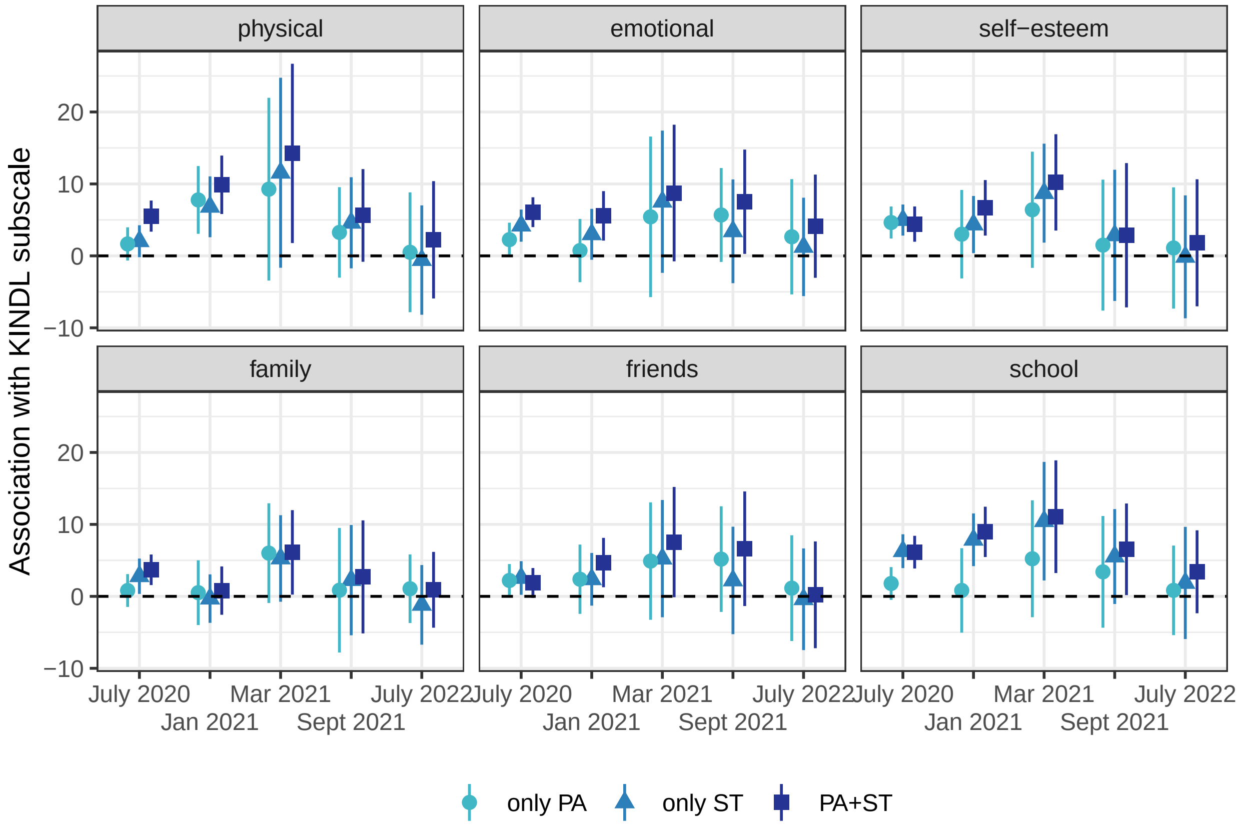

Figure S2: Model results for association of HRQOL subscales with adherence to physical activity and screen time recommendations (compared to reference category of not adhering to either recommendation).

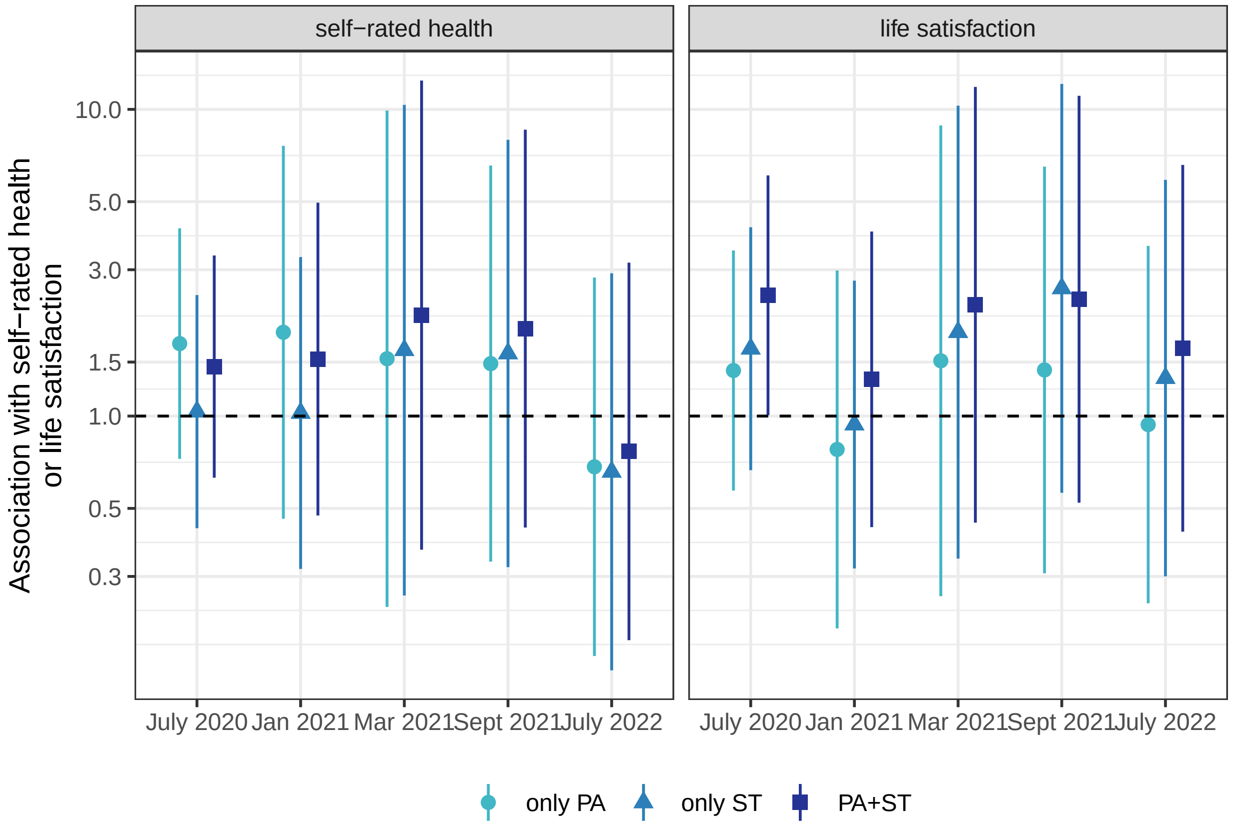

Figure S3: Model results for association of self-rated health and life satisfaction with adherence to physical activity and screen time recommendations (odds ratio (95% confidence interval))

Table S3: Observed HRQOL by adherence to physical activity and screen time recommendations (median [intraquartile range])

| timepoint | n | neither | only PA | only ST | PA+ST |
|---|---|---|---|---|---|
| July 2020 | 1769 | 76.0 [67.7, 82.3] | 78.1 [71.9, 84.4] | 80.2 [74.0, 85.4] | 82.3 [77.1, 87.5] |
| Jan 2021 | 1572 | 71.9 [63.5, 78.1] | 72.9 [66.7, 81.2] | 77.1 [69.8, 82.3] | 80.2 [74.0, 85.4] |
| Mar 2021 | 1494 | 70.8 [62.5, 78.1] | 74.0 [66.7, 81.2] | 77.1 [71.2, 83.3] | 80.2 [72.9, 85.4] |
| Sept 2021 | 922 | 71.9 [63.5, 79.2] | 75.0 [67.7, 81.2] | 75.0 [68.8, 81.2] | 80.2 [74.0, 85.4] |
| July 2022 | 809 | 70.8 [64.6, 78.9] | 76.0 [68.9, 82.3] | 76.1 [70.8, 81.2] | 79.2 [72.9, 85.1] |

Table S4: Model results for association of HRQOL with adherence to physical activity and screen time recommendations (coefficient (95% confidence interval))

| item | timepoint | neither | only PA | only ST | PA+ST |
|---|---|---|---|---|---|
| total | July 2020 | (reference) | 2.2 (0.4-4.0) | 3.8 (1.9-5.7) | 4.6 (2.8-6.5) |
| total | Jan 2021 | (reference) | 2.0 (-1.8-5.8) | 3.5 (0.4-6.7) | 5.5 (2.4-8.5) |
| total | Mar 2021 | (reference) | 6.2 (-0.5-12.9) | 8.1 (1.1-15.2) | 9.7 (3.0-16.3) |
| total | Sept 2021 | (reference) | 3.2 (-2.3-8.8) | 3.4 (-2.0-8.8) | 5.2 (-0.7-11.0) |
| total | July 2022 | (reference) | 1.7 (-2.3-5.6) | 0.5 (-3.4-4.4) | 2.4 (-1.3-6.1) |

Table S5: Model results for association of HRQOL physical, emotional and self-esteem subscales with adherence to physical activity and screen time recommendations (coefficient (95% confidence interval))

| item | timepoint | neither | only PA | only ST | PA+ST |
|---|---|---|---|---|---|
| physical | July 2020 | (reference) | 1.7 (-0.6-4.0) | 2.0 (-0.2-4.3) | 5.5 (3.4-7.7) |
| physical | Jan 2021 | (reference) | 7.8 (3.1-12.5) | 6.8 (2.6-11.0) | 9.9 (5.8-13.9) |
| physical | Mar 2021 | (reference) | 9.3 (-3.4-22.0) | 11.6 (-1.6-24.8) | 14.2 (1.8-26.7) |
| physical | Sept 2021 | (reference) | 3.3 (-3.0-9.5) | 4.6 (-1.7-10.9) | 5.6 (-0.8-12.1) |
| physical | July 2022 | (reference) | 0.5 (-7.8-8.8) | -0.6 (-8.2-7.0) | 2.2 (-5.9-10.4) |
| emotional | July 2020 | (reference) | 2.3 (-0.1-4.6) | 4.2 (2.0-6.4) | 6.1 (4.0-8.1) |
| emotional | Jan 2021 | (reference) | 0.7 (-3.7-5.1) | 3.0 (-0.5-6.5) | 5.6 (2.1-9.0) |
| emotional | Mar 2021 | (reference) | 5.4 (-5.7-16.6) | 7.5 (-2.4-17.4) | 8.7 (-0.8-18.2) |
| emotional | Sept 2021 | (reference) | 5.7 (-0.8-12.2) | 3.4 (-3.8-10.6) | 7.5 (0.3-14.8) |
| emotional | July 2022 | (reference) | 2.7 (-5.4-10.7) | 1.2 (-5.6-8.1) | 4.1 (-3.0-11.3) |
| self-esteem | July 2020 | (reference) | 4.6 (2.4-6.9) | 5.0 (2.8-7.1) | 4.4 (2.0-6.9) |
| self-esteem | Jan 2021 | (reference) | 3.0 (-3.1-9.2) | 4.4 (0.4-8.3) | 6.7 (2.8-10.5) |
| self-esteem | Mar 2021 | (reference) | 6.4 (-1.7-14.5) | 8.7 (1.8-15.6) | 10.2 (3.5-16.9) |
| self-esteem | Sept 2021 | (reference) | 1.5 (-7.6-10.6) | 2.9 (-6.3-12.0) | 2.9 (-7.2-12.9) |
| self-esteem | July 2022 | (reference) | 1.1 (-7.3-9.5) | -0.1 (-8.7-8.4) | 1.8 (-7.0-10.6) |

Table S6: Model results for association of HRQOL family, friends and school subscales with adherence to physical activity and screen time recommendations (coefficient (95% confidence interval))

| item | timepoint | neither | only PA | only ST | PA+ST |
|---|---|---|---|---|---|
| family | July 2020 | (reference) | 0.8 (-1.5-3.1) | 2.8 (0.3-5.3) | 3.7 (1.6-5.8) |
| family | Jan 2021 | (reference) | 0.5 (-4.0-5.0) | -0.3 (-3.7-3.1) | 0.8 (-2.6-4.2) |
| family | Mar 2021 | (reference) | 6.0 (-0.9-12.9) | 5.3 (-0.7-11.3) | 6.1 (0.2-12.0) |
| family | Sept 2021 | (reference) | 0.9 (-7.8-9.5) | 2.3 (-5.4-9.9) | 2.7 (-5.1-10.6) |
| family | July 2022 | (reference) | 1.1 (-3.7-5.8) | -1.2 (-6.7-4.4) | 0.9 (-4.4-6.2) |
| friends | July 2020 | (reference) | 2.2 (-0.1-4.5) | 2.6 (0.2-4.9) | 1.9 (-0.2-3.9) |
| friends | Jan 2021 | (reference) | 2.4 (-2.4-7.2) | 2.4 (-1.3-6.0) | 4.7 (1.3-8.1) |
| friends | Mar 2021 | (reference) | 4.9 (-3.3-13.1) | 5.2 (-2.9-13.4) | 7.5 (-0.1-15.2) |
| friends | Sept 2021 | (reference) | 5.2 (-2.2-12.5) | 2.2 (-5.3-9.7) | 6.6 (-1.3-14.6) |
| friends | July 2022 | (reference) | 1.1 (-6.2-8.5) | -0.4 (-7.5-6.7) | 0.2 (-7.2-7.6) |
| school | July 2020 | (reference) | 1.8 (-0.5-4.1) | 6.3 (3.9-8.6) | 6.1 (3.9-8.4) |
| school | Jan 2021 | (reference) | 0.8 (-5.0-6.7) | 7.9 (4.2-11.5) | 9.0 (5.5-12.5) |
| school | Mar 2021 | (reference) | 5.2 (-2.9-13.3) | 10.4 (2.2-18.7) | 11.1 (3.2-18.9) |
| school | Sept 2021 | (reference) | 3.4 (-4.4-11.2) | 5.5 (-1.1-12.1) | 6.5 (0.2-12.9) |
| school | July 2022 | (reference) | 0.8 (-5.4-7.1) | 1.9 (-5.9-9.7) | 3.4 (-2.3-9.2) |

Table S7: Model results for association of self-rated health and life satisfaction with adherence to physical activity and screen time recommendations (odds ratio (95% confidence interval))

| item | timepoint | neither | only PA | only ST | PA+ST |
|---|---|---|---|---|---|
| self-rated health | July 2020 | (reference) | 1.72 (5.60-5.60) | 1.03 (2.81-2.81) | 1.45 (4.26-4.26) |
| self-rated health | Jan 2021 | (reference) | 1.88 (6.53-6.53) | 1.02 (2.78-2.78) | 1.53 (4.64-4.64) |
| self-rated health | Mar 2021 | (reference) | 1.54 (4.66-4.66) | 1.64 (5.15-5.15) | 2.13 (8.45-8.45) |
| self-rated health | Sept 2021 | (reference) | 1.48 (4.41-4.41) | 1.60 (4.95-4.95) | 1.93 (6.88-6.88) |
| self-rated health | July 2022 | (reference) | 0.68 (1.98-1.98) | 0.66 (1.93-1.93) | 0.77 (2.15-2.15) |
| life satisfaction | July 2020 | (reference) | 1.41 (4.09-4.09) | 1.66 (5.25-5.25) | 2.48 (11.90-11.90) |
| life satisfaction | Jan 2021 | (reference) | 0.78 (2.18-2.18) | 0.94 (2.56-2.56) | 1.32 (3.73-3.73) |
| life satisfaction | Mar 2021 | (reference) | 1.51 (4.55-4.55) | 1.88 (6.54-6.54) | 2.31 (10.03-10.03) |
| life satisfaction | Sept 2021 | (reference) | 1.41 (4.11-4.11) | 2.61 (13.58-13.58) | 2.40 (11.06-11.06) |
| life satisfaction | July 2022 | (reference) | 0.94 (2.55-2.55) | 1.33 (3.78-3.78) | 1.66 (5.28-5.28) |

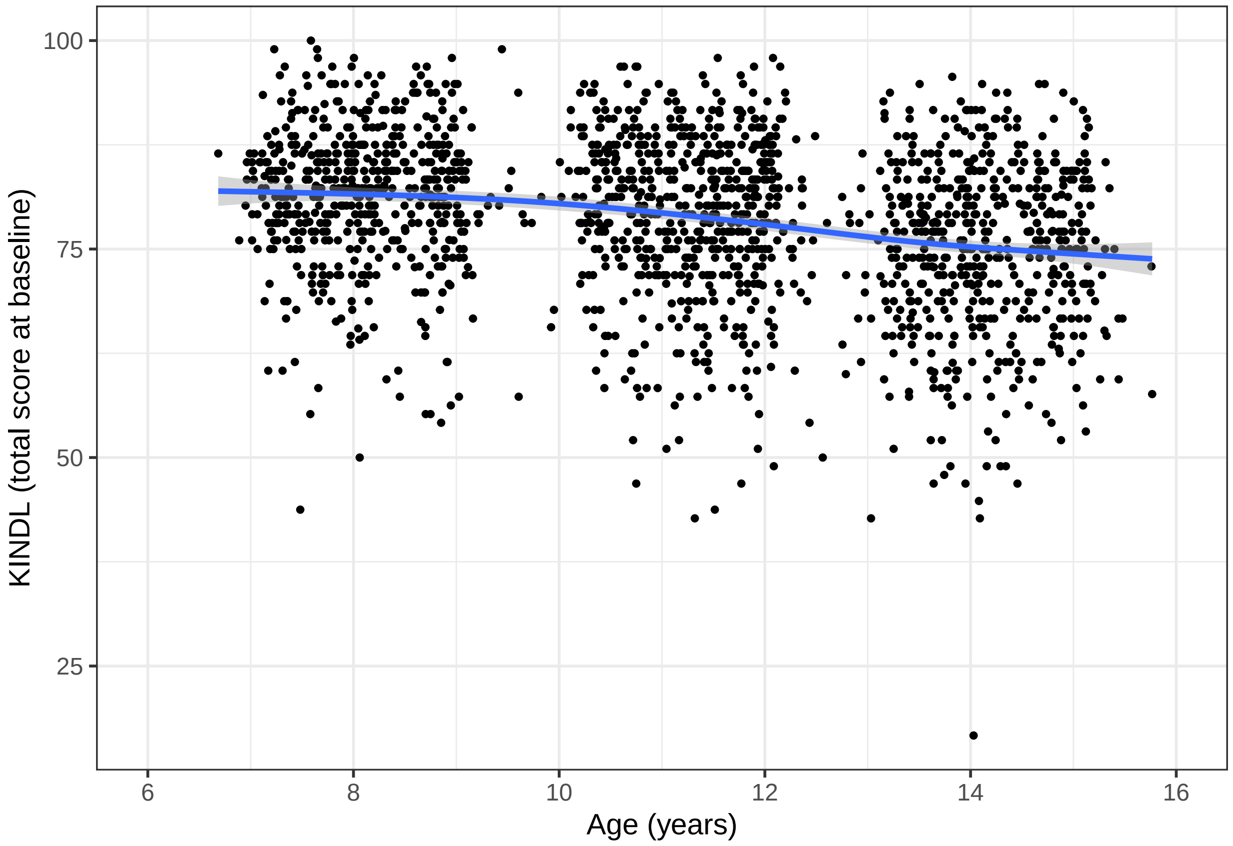

Figure S4: HRQOL by age at baseline

Table S8: Age-stratified model results for association of HRQOL with adherence to physical activity and screen time recommendations (coefficient (95% confidence interval))

| grp | item | timepoint | neither | only PA | only ST | PA+ST |
|---|---|---|---|---|---|---|
| primary | total | July 2020 | (reference) | 2.1 (-0.1-4.2) | 3.0 (1.0-5.0) | 4.8 (2.8-6.7) |
| primary | total | Jun 2021 | (reference) | 2.4 (-3.6-8.4) | 4.1 (0.2-8.0) | 4.8 (1.0-8.6) |
| primary | total | Mar 2021 | (reference) | 0.2 (-8.3-8.7) | 3.9 (-4.0-11.8) | 5.2 (-2.4-12.8) |
| primary | total | Sept 2021 | (reference) | 2.3 (-4.6-9.2) | 2.6 (-3.3-8.5) | 3.8 (-1.7-9.4) |
| primary | total | July 2022 | (reference) | 2.3 (-3.0-7.6) | 0.7 (-4.1-5.5) | 3.3 (-1.8-8.4) |
| secondary | total | July 2020 | (reference) | 1.9 (-0.3-4.1) | 4.3 (1.7-6.8) | 5.6 (3.2-8.1) |
| secondary | total | Jun 2021 | (reference) | 0.1 (-3.0-3.2) | 0.6 (-2.2-3.4) | 5.0 (2.0-7.9) |
| secondary | total | Mar 2021 | (reference) | 14.0 (2.2-25.7) | 13.8 (2.3-25.4) | 15.3 (3.6-27.0) |
| secondary | total | Sept 2021 | (reference) | 2.0 (-3.2-7.2) | 1.1 (-4.9-7.0) | 4.1 (-2.4-10.6) |
| secondary | total | July 2022 | (reference) | 1.3 (-3.0-5.6) | 0.6 (-5.2-6.4) | 1.1 (-3.8-5.9) |
